# Diagnostic accuracy of tongue swab testing in persons with sputum Xpert Ultra Trace results

**DOI:** 10.1101/2025.07.15.25331566

**Authors:** Adrienne E. Shapiro, Ronit R Dalmat, Job Mukwatamundu, Caleb Kamoga, M William Ngwane, Amy Steadman, Elvira Budiawan, Gabrielle Stein, Annet Nalutaaya, James Mukiibi, Mariam Nantale, Patrick Biché, Caitlin Visek, Joowhan Sung, Zanele Magcaba, Nompumelelo Ngcobo, Jennifer F. Morton, Meena Lenn, Samantha Aucock, Robin Draper, Amanda Ganguloo, Douglas Wilson, Achilles Katamba, Emily A Kendall, Paul K Drain

## Abstract

**Background:** Molecular amplification of tongue swab samples is a non-sputum-based investigational approach to diagnose pulmonary tuberculosis (pTB). An improved manual qPCR method for tongue swabs recently achieved >90% sensitivity overall in diagnosing TB, compared to a sputum microbiologic reference standard. Performance characteristics in persons with low-positive results on sputum molecular tests are unknown.

**Methods:** Adults in South Africa and Uganda with sputum Xpert MTB/RIF Ultra Trace (TR+) results were recruited for confirmatory evaluation and follow-up. They underwent symptom evaluation, examination, chest X-ray, further sputum testing (repeat Xpert Ultra and two solid and liquid mycobacterial cultures), and two tongue swabs. Tongue swabs were tested using qPCR amplification of the IS6110 gene. A single copy detected on >=1 swab was considered TB-positive. TR+ persons not diagnosed with TB at baseline were re-evaluated at 1 and 3 months. We determined the sensitivity and specificity of tongue swabs against TB culture alone, a microbiologic reference standard (MRS: any positive result from Xpert Ultra or TB culture) and a composite reference standard (CRS: a clinical recommendation for TB treatment or any positive culture) at baseline.

**Results:** 225 enrolled TR+ participants (115 (51%) women, median age 38 [IQR 30-47], 130 (58%) people living with HIV (PWH)) provided at least 1 tongue swab at baseline. With a culture reference standard, 45 (20%) were positive for TB at baseline testing; 58 (26%) were positive for TB by MRS and 83 (37%) by CRS. Sensitivity and specificity of tongue swabs against culture were 25% [95% CI 13-40%] and 94% [90-97%], vs. MRS were 25% [95% CI 14-38%] and 96% [91-98%], and vs. CRS were 16% [9-26%] and 94% [89-98%].

**Conclusion:** Tongue swabs had low sensitivity and moderately high specificity for TB in persons with a Trace Xpert Ultra result. Tongue swabs have limited value for diagnosing people with low-positive molecular test results of uncertain clinical significance.

## Introduction

A key barrier to decreasing tuberculosis (TB) morbidity and mortality globally is the lack of sensitive diagnostic tools which do not rely on sputum sampling. TB is the leading cause of infectious diseases mortality worldwide, causing illness in 10.6 million people and death in over one million people worldwide in 2024.^1^ A significant diagnostic gap persists between estimated and microbiologically diagnosed cases of pulmonary TB.

The current WHO-recommended initial diagnostic test for a person with signs or symptoms of tuberculosis is a low-complexity nucleic acid amplification test (lcNAAT) performed on a sputum specimen.^2^ The Xpert MTB/RIF Ultra (“Xpert Ultra”) cartridge-based test is the leading lcNAAT test available globally, and the test returns a semiquantitative result of “High,” “Medium,” “Low,” “Very Low” or “Trace” when *Mycobacterium tuberculosis* DNA is detected in the patient sample.^3^ There is currently clinical uncertainty about the significance of a “Trace” (TR+) sputum result, which may represent true TB disease, but could also represent residual bacteria from treated disease or low-bacterial-burden disease which could either progress to higher TB burden disease or may resolve without intervention.^4^

Tongue swabs are a promising new diagnostic approach for pulmonary TB, using a swab to collect TB bacterial and genetic matter that is deposited on the tongue from the respiratory tract, and using a molecular assay to amplify targets from the TB genome in the tongue swab sample. As a non-sputum-based testing approach, tongue swabs are an attractive approach to increase yield of sample collection and TB diagnosis in persons who do not easily produce sputum, including persons with asymptomatic TB, children, and persons with advanced HIV disease.^5^ Ongoing product development and optimization of tongue swab analytic approaches and devices have improved testing options since a systematic review in 2024 identified sensitivities of diverse tongue swab tests ranging from 36%-91% in adults.^6^ A tongue swab diagnostic test using a manual qPCR analysis produced a sensitivity of 92.6% and specificity of 99.1% compared to a microbiologic reference standard in 397 adults in Uganda evaluated for TB.^7^ This and other studies included only small numbers of individuals with Trace sputum Xpert results and included only limited investigations to clarify those individuals’ true TB status; however, sensitivity was directly related to Xpert Ultra semi-quantitative sputum result, suggesting tongue swabs may have limited diagnostic value in persons with low bacterial sputum loads.^8^

As part of a prospective cohort study undertaken to determine the clinical and microbiological significance of the Xpert Ultra Trace result (TR+), we evaluated the diagnostic test accuracy of the tongue swabs qPCR assay in a population of adults and adolescents undergoing confirmatory TB diagnostic evaluation after receiving a TR+ result in routine clinical care. This focused evaluation was done to assess the utility of tongue swabs in diagnosing TB in persons with low bacterial load sputum.

## Methods

### Study design, participants, and procedures

Adults and adolescents aged 16 years and older in Uganda and South Africa with Trace Xpert MTB/RIF Ultra results on sputum (TR+) obtained in routine clinical settings enrolled in a prospective cohort study. Study procedures for this cohort investigating the clinical and microbiological significance of a Trace result are previously described.^9^ Briefly, eligible participants had a TR+ sputum result, had not yet initiated TB treatment, did not require hospitalization, and had not received TB treatment within the previous 3 months. TR+ participants were enrolled between 2022-2024.

At the study baseline visit conducted within 14 days of the TR+ result, participants were systematically evaluated for TB disease with investigations including symptom screen, physical exam, digital chest X-ray (CXR), repeat sputum collection for testing with Xpert Ultra and TB culture (liquid and solid culture on 2 specimens), and C-reactive protein. TR+ participants with HIV also had CD4+T-cell counts measured and provided a urine sample for urine LAM testing. With review of clinical data, study physician panels at each site or the referring clinician (Uganda site) could recommend initiation of TB treatment or continued deferral.

After clinical review of the participant and data, participants who were not recommended to start TB treatment were recalled to the study clinic for repeat sputum assessments, clinical evaluations, and CXR at one month and three months.

### Tongue swab sample and control collection

To investigate the diagnostic utility of tongue swabs in this population of TR+ individuals, participants were asked to provide at least two tongue swab samples. Tongue swabs were collected prior to sputum production at the baseline visit. A study clinician collected up to 3 Copan *FLOQswab* spun polyester swab samples of the tongue surface according to a standardized protocol.^10^ Tongue swabs were eluted into 500ul of TE buffer in a cryovial, sealed, and transported to the study lab in a cooler box at 2-6C for testing. Tongue swabs not tested immediately were stored at −80C until analysis.

To detect possible contamination as a source of positive results in the nucleic acid amplification step, “air controls” were obtained at each site by exposing a sterile swab to the air in the clinical collection site.. Air controls were obtained weekly at the South Africa site and for each participant at Uganda sites and were processed in an identical manner alongside the participant samples.

### Sample preparation and index test analysis

Tongue swab samples were analyzed at each site using an investigational manual quantitative PCR assay developed by Steadman and colleagues.^7^ Briefly, tongue swab samples were thawed (if frozen), vortexed, and heat-inactivated for 10 minutes at 95°C. After bead-beater cycles to lyse cells, the lysate was applied to a PCR plate and processed on a thermal cycler with oligonucleotide sequences designed to amplify the *IS6110* insertion sequence of *M. tuberculosis*. Detection of a single copy of the target sequence in any tongue swab was considered positive for detection of tuberculosis in that participant. Because the tongue swab approach is investigational and not yet approved for use in clinical practice, no tongue swab results were disclosed to the participant or treating clinicians.Tongue swab processing and qPCR was conducted without the knowledge of the results of reference testing of the participant.

### Reference definitions for TB diagnosis at a given timepoint

Three TB diagnostic reference standards were considered for the TR+ population in evaluating tongue swabs as an index test: 1) ***TB culture***, defined as at least one sputum TB culture yielding a positive result for *M. tuberculosis* complex. Negative or missing cultures were considered negative. Missing cultures in a participant who was recommended to receive TB treatment were excluded. 2) ***Microbiologic reference standard (MRS)*** *positive*, defined as at least one of sputum Xpert Ultra positive (Mtb complex detected, not Trace) or sputum culture positive; negative if sputum Xpert Ultra and all sputum cultures were negative or missing. Persons with a missing MRS result who were treated were excluded. 3) ***Composite reference standard (CRS)*** positive, defined as at least one of sputum culture positive or a clinical recommendation to initiate TB treatment.

### Data analysis

We calculated sensitivity, specificity, and 95% confidence intervals for the index test (tongue swabs analyzed by qPCR) against each of three reference standards. In stratified analyses, we calculated sensitivity and specificity for tongue swabs by HIV status of the TR+ participant, by prior TB status of the TR+ participant, and by baseline (repeat) Xpert Ultra sputum results of the TR+ participant.

### Ethical considerations

The study was approved by the Makerere University School of Public Health Research and Ethics Committee, the University of KwaZulu-Natal Biomedical Research Ethics Committee, and the institutional review boards at the University of Washington and Johns Hopkins University.

## Results

Across both sites, 225 adults with initial sputum TR+ results provided at least one tongue swab (up to 2 swabs) for testing at the baseline visit (**Table 1**). Male and female participants were similarly represented (110 male, 49%), with median age 38 years (IQR 30, 47). Over half of the TR+ participants had HIV (130, 58%). At baseline screening, 199 (88%) of the cohort reported at least one sign or symptom of TB, and 85 (38%) had been diagnosed with TB previously.

**Table 1.**
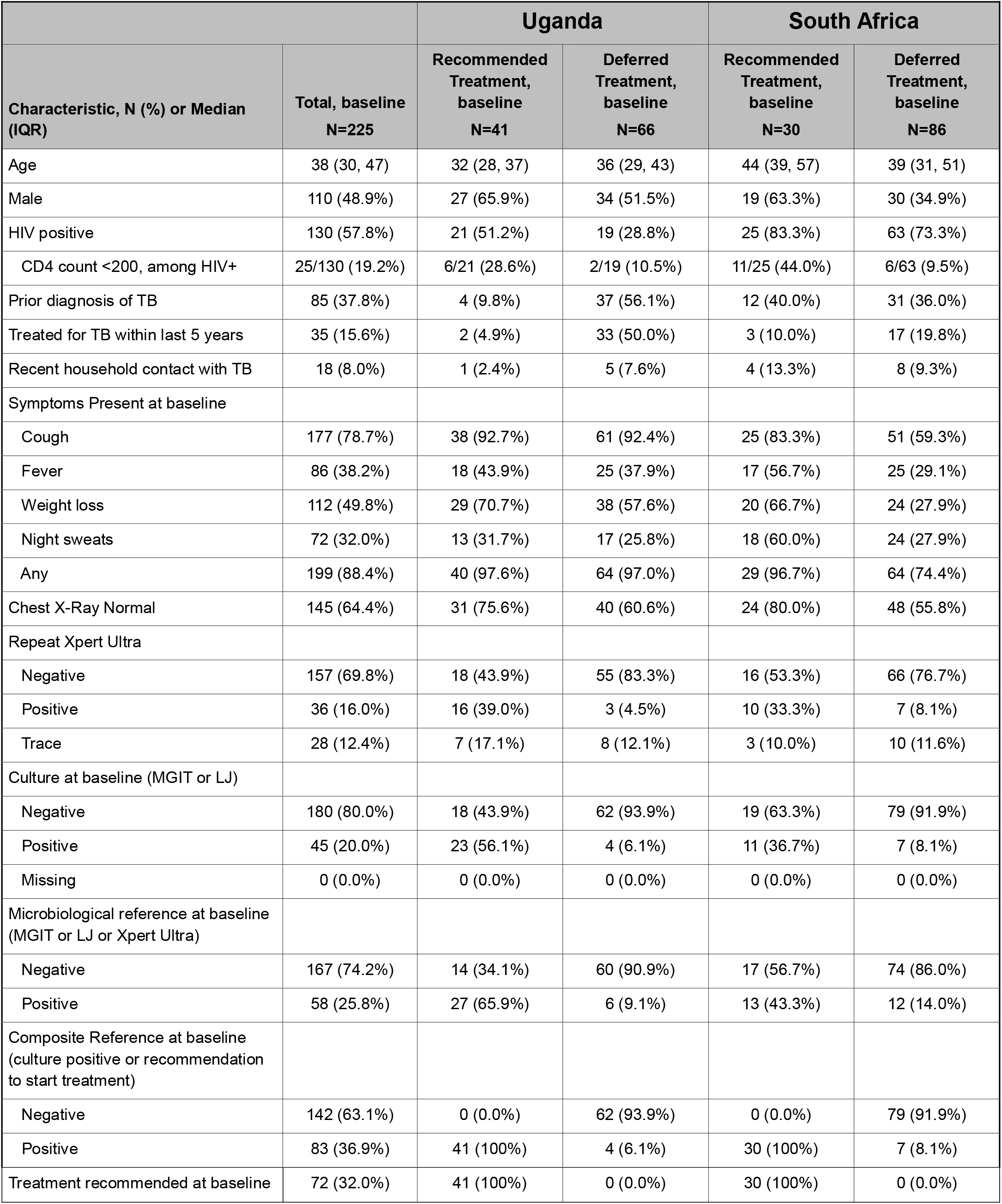
Characteristics of persons with TR+ sputum evaluated for TB using tongue swabs.

|  |  | Uganda |  | South Africa |  |
| --- | --- | --- | --- | --- | --- |
| Characteristic, N (%) or Median (IQR) | Total, baseline<br>N=225 | Recommended<br>Treatment,<br>baseline<br>N=41 | Deferred<br>Treatment,<br>baseline<br>N=66 | Recommended<br>Treatment,<br>baseline<br>N=30 | Deferred<br>Treatment,<br>baseline<br>N=86 |
| Age | 38 (30, 47) | 32 (28, 37) | 36 (29, 43) | 44 (39, 57) | 39 (31, 51) |
| Male | 110 (48.9%) | 27 (65.9%) | 34 (51.5%) | 19 (63.3%) | 30 (34.9%) |
| HIV positive | 130 (57.8%) | 21 (51.2%) | 19 (28.8%) | 25 (83.3%) | 63 (73.3%) |
| CD4 count <200, among HIV+ | 25/130 (19.2%) | 6/21 (28.6%) | 2/19 (10.5%) | 11/25 (44.0%) | 6/63 (9.5%) |
| Prior diagnosis of TB | 85 (37.8%) | 4 (9.8%) | 37 (56.1%) | 12 (40.0%) | 31 (36.0%) |
| Treated for TB within last 5 years | 35 (15.6%) | 2 (4.9%) | 33 (50.0%) | 3 (10.0%) | 17 (19.8%) |
| Recent household contact with TB | 18 (8.0%) | 1 (2.4%) | 5 (7.6%) | 4 (13.3%) | 8 (9.3%) |
| Symptoms Present at baseline |  |  |  |  |  |
| Cough | 177 (78.7%) | 38 (92.7%) | 61 (92.4%) | 25 (83.3%) | 51 (59.3%) |
| Fever | 86 (38.2%) | 18 (43.9%) | 25 (37.9%) | 17 (56.7%) | 25 (29.1%) |
| Weight loss | 112 (49.8%) | 29 (70.7%) | 38 (57.6%) | 20 (66.7%) | 24 (27.9%) |
| Night sweats | 72 (32.0%) | 13 (31.7%) | 17 (25.8%) | 18 (60.0%) | 24 (27.9%) |
| Any | 199 (88.4%) | 40 (97.6%) | 64 (97.0%) | 29 (96.7%) | 64 (74.4%) |
| Chest X-Ray Normal | 145 (64.4%) | 31 (75.6%) | 40 (60.6%) | 24 (80.0%) | 48 (55.8%) |
| Repeat Xpert Ultra |  |  |  |  |  |
| Negative | 157 (69.8%) | 18 (43.9%) | 55 (83.3%) | 16 (53.3%) | 66 (76.7%) |
| Positive | 36 (16.0%) | 16 (39.0%) | 3 (4.5%) | 10 (33.3%) | 7 (8.1%) |
| Trace | 28 (12.4%) | 7 (17.1%) | 8 (12.1%) | 3 (10.0%) | 10 (11.6%) |
| Culture at baseline (MGIT or LJ) |  |  |  |  |  |
| Negative | 180 (80.0%) | 18 (43.9%) | 62 (93.9%) | 19 (63.3%) | 79 (91.9%) |
| Positive | 45 (20.0%) | 23 (56.1%) | 4 (6.1%) | 11 (36.7%) | 7 (8.1%) |
| Missing | 0 (0.0%) | 0 (0.0%) | 0 (0.0%) | 0 (0.0%) | 0 (0.0%) |
| Microbiological reference at baseline (MGIT or LJ or Xpert Ultra) |  |  |  |  |  |
| Negative | 167 (74.2%) | 14 (34.1%) | 60 (90.9%) | 17 (56.7%) | 74 (86.0%) |
| Positive | 58 (25.8%) | 27 (65.9%) | 6 (9.1%) | 13 (43.3%) | 12 (14.0%) |
| Composite Reference at baseline (culture positive or recommendation to start treatment) |  |  |  |  |  |
| Negative | 142 (63.1%) | 0 (0.0%) | 62 (93.9%) | 0 (0.0%) | 79 (91.9%) |
| Positive | 83 (36.9%) | 41 (100%) | 4 (6.1%) | 30 (100%) | 7 (8.1%) |
| Treatment recommended at baseline | 72 (32.0%) | 41 (100%) | 0 (0.0%) | 30 (100%) | 0 (0.0%) |

At the end of baseline evaluations, 45 (20%) of TR+ participants were positive for TB by culture, 58 (26%) were positive for TB by MRS and 83 (37%) were positive for TB by CRS. Complete baseline data was available for evaluation for 217 participants. Four participants had indeterminate tongue swab results, and one participant had tongue swab data excluded due to a positive air control swab for that person.

The overall sensitivity of tongue swabs to diagnose TB in persons with TR+ sputum ranged from 16% (95% CI) to 25% (95% CI), depending on the reference standard used (**Table 2**). Specificity ranged from 99% (95% CI 93, 100)) to 100% (95% CI 100,100). There was little difference between sensitivity and specificity seen across the two study sites. Analyses stratified by HIV status (**Table 3**) and prior TB status of participants (**Table 4**) showed similarly consistent low sensitivity, regardless of HIV status and prior TB status. The sensitivity of tongue swabs relative to culture was modestly higher in persons with a baseline (repeat) sputum Xpert Ultra result showing a higher semiquantitative value (**Table 5**), although detection of a meaningful trend is limited by very small numbers of people with high or medium Xpert Ultra results on repeat testing.

**Table 2.**
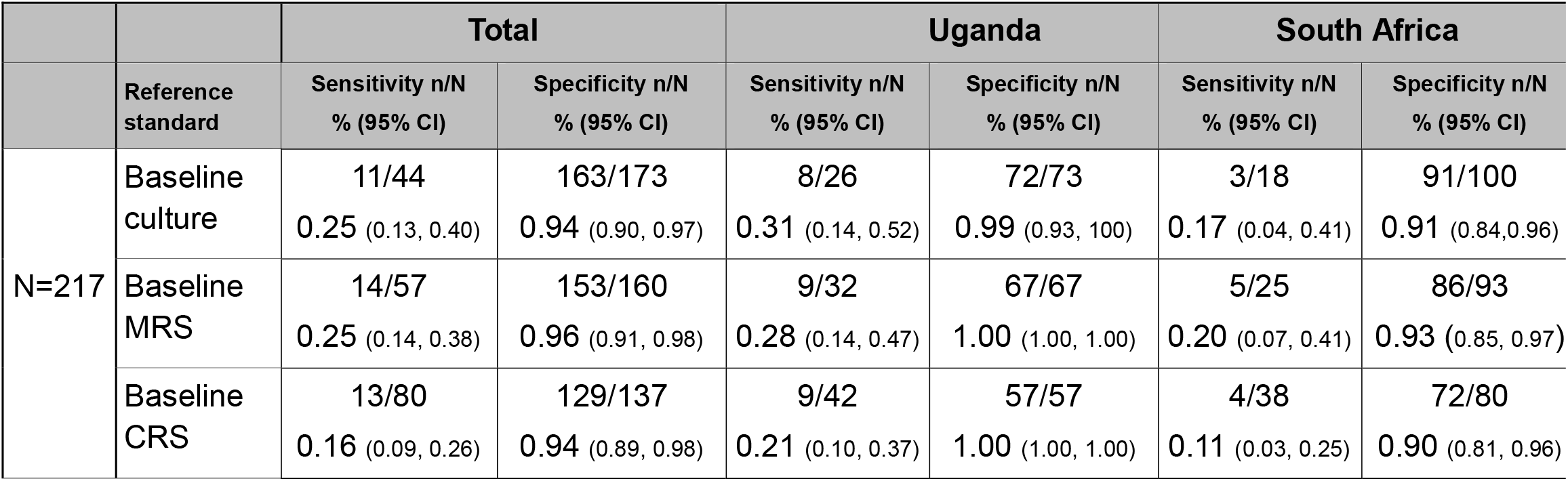
Overall sensitivity and specificity of tongue swabs in persons with initial TR+ result, against three reference standards.

|  |  | Total |  | Uganda |  | South Africa |  |
| --- | --- | --- | --- | --- | --- | --- | --- |
|  | Reference standard | Sensitivity n/N<br>% (95% CI) | Specificity n/N<br>% (95% CI) | Sensitivity n/N<br>% (95% CI) | Specificity n/N<br>% (95% CI) | Sensitivity n/N<br>% (95% CI) | Specificity n/N<br>% (95% CI) |
| N=217 | Baseline culture | 11/44<br>0.25 (0.13, 0.40) | 163/173<br>0.94 (0.90, 0.97) | 8/26<br>0.31 (0.14, 0.52) | 72/73<br>0.99 (0.93, 1.00) | 3/18<br>0.17 (0.04, 0.41) | 91/100<br>0.91 (0.84, 0.96) |
|  | Baseline MRS | 14/57<br>0.25 (0.14, 0.38) | 153/160<br>0.96 (0.91, 0.98) | 9/32<br>0.28 (0.14, 0.47) | 67/67<br>1.00 (1.00, 1.00) | 5/25<br>0.20 (0.07, 0.41) | 86/93<br>0.93 (0.85, 0.97) |
|  | Baseline CRS | 13/80<br>0.16 (0.09, 0.26) | 129/137<br>0.94 (0.89, 0.98) | 9/42<br>0.21 (0.10, 0.37) | 57/57<br>1.00 (1.00, 1.00) | 4/38<br>0.11 (0.03, 0.25) | 72/80<br>0.90 (0.81, 0.96) |

**Table 3.**
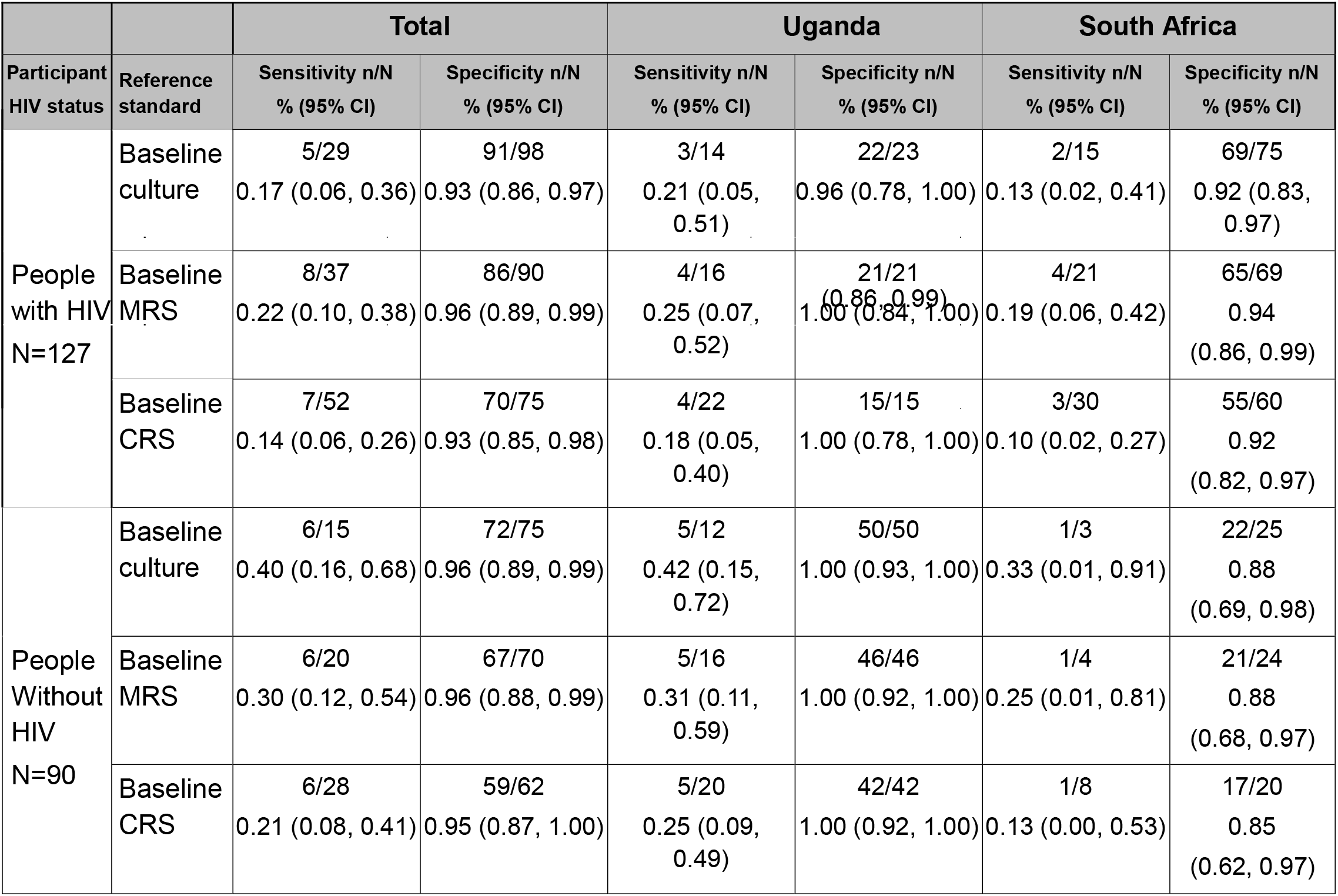
Diagnostic accuracy (sensitivity and specificity) of tongue swabs for pulmonary tuberculosis in TR+ persons, against 3 reference standards, stratified by HIV status.

**Table 4.**
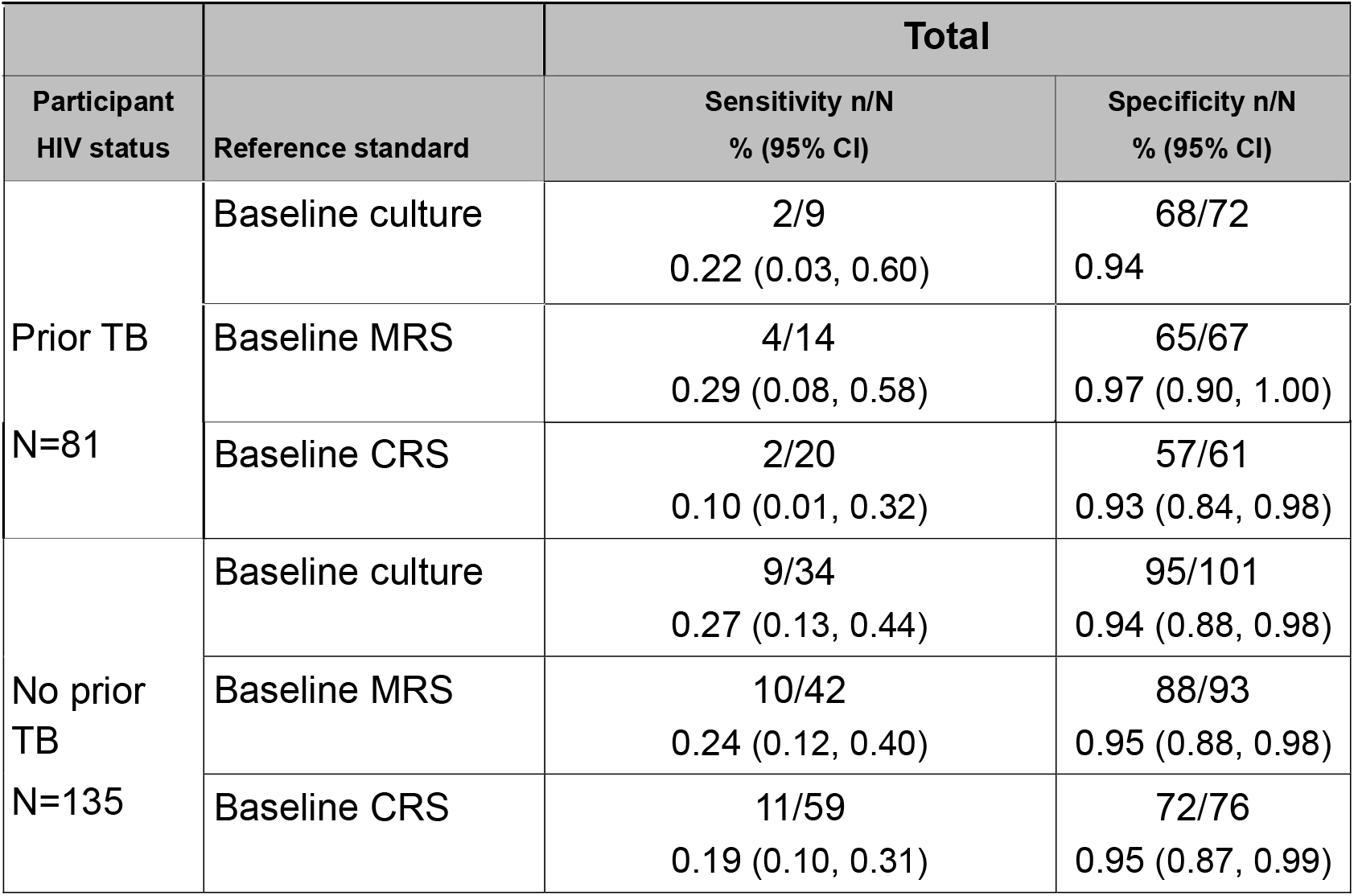
Diagnostic accuracy (sensitivity and specificity) of tongue swabs for pulmonary tuberculosis in TR+ persons, against 3 reference standards, stratified by prior TB status.

**Table 5.** Sensitivity and specificity of tongue swabs for TB diagnosis, stratified by baseline repeat Xpert Ultra sputum result. Reference standard: TB culture.

|  |  | Total |  |
| --- | --- | --- | --- |
| Ref: baseline culture | Baseline GXP Result | Sensitivity n/N<br>% (95% CI) | Specificity n/N<br>% (95% CI) |
|  | High or medium | 1/1<br>1.00 (0.02, 1.00) | N/A |
|  | Low or very low | 7/21<br>0.33 (0.15, 0.57) | 10/13<br>0.77 (0.46, 0.95) |
|  | Trace | 1/10<br>0.10 (0.00, 0.45) | 18/18<br>1.00 (0.82, 1.00) |
|  | Negative | 2/11<br>0.18 (0.02, 0.52) | 134/138<br>0.97 (0.93, 0.99) |

## Discussion

In a cohort of adults and adolescents in South Africa and Uganda with sputum Xpert Ultra Trace results undergoing intensive investigation for TB, we found that tongue swabs analyzed with a manual qPCR protocol had very low sensitivity and moderately high specificity to diagnose TB. Tongue swab sensitivity was consistently less than 30%, regardless of reference standard for TB used (TB culture, MRS, or CRS), HIV status of the participant, or prior TB status of the participant. Notably, this same qPCR method with tongue swabs demonstrated high sensitivity, in excess of 90%, for diagnosing TB in a large cohort of Ugandan adults *not* stratified by sputum bacterial quantity, suggesting the specific limitation of the tongue swab diagnostic approach to persons with low sputum TB burden.^7^

Even with suboptimal sensitivity, tongue swabs may provide important diagnostic options for detecting TB in certain settings, including persons who cannot readily produce sputum, as shown recently for field sample collection in a household contact-tracing study.^11^ Our study of tongue swabs after a Trace sputum Xpert result also found that they offered some incremental detection of culture-positive TB among people in whom repeat sputum testing would have yielded a false negative result. Thus, tongue swabs may have some diagnostic role even in persons with low bacterial burden, potentially even in limited settings where sputum collection is possible. However, the overwhelming finding that tongue swabs were usually negative in persons with TR+ sputum, and correlated only weakly with other investigations to clarify these individuals’ true TB status, highlights paucibacillary TB as a significant limitation of tongue swabs as a diagnostic strategy. As the potential role for tongue swabs for TB diagnosis continues to be defined, our findings underscore the need to incorporate semiquantitative sputum bacterial levels order to meaningfully interpret diagnostic test accuracy.

In conclusion, tongue swabs analyzed with a qPCR assay had low sensitivity and moderately high specificity for TB in persons with a Trace Xpert Ultra sputum result indicating low sputum bacterial burden. Tongue swabs have limited value for diagnosing people with low-positive molecular sputum test results.

## Data Availability

The deidentified dataset used for this study and a data dictionary are being uploaded to a controlled access data archive, per
requirements of the institutional review board that reviewed and approved this study. The upload process is under review and this
preprint will be updated with a link to the data as soon as available. At this time, all data produced in the present study are available
upon reasonable request to the authors.

## Acknowledgements

This work would not be possible without the support of the staff of Harry Gwala Regional Hospital and referring primary health care clinics, the KwaZulu-Natal Department of Health, the South African National Health Laboratory Service, the TURN-TB research team and physician consultants, and the clinical and laboratory staff at Kitebi Health Centre, Kisugu Health Centre, Kisenyi Health Centre, Kawaala Health Centre, Kiswa Health Centre, Naguru Hospital, and Mulago Hospital Wards 5&6.

